# Trajectories of anxiety and depressive symptoms during enforced isolation due to COVID-19: longitudinal analyses of 36,520 adults in England

**DOI:** 10.1101/2020.06.03.20120923

**Authors:** Daisy Fancourt, Andrew Steptoe, Feifei Bu

## Abstract

**Background:** There is currently major concern about the impact of the global COVID-19 outbreak on mental health. A number of studies suggest that mental health deteriorated in many countries prior to and during enforced isolation (“lockdown”), but it remains unknown how mental health has changed week by week over the course of the COVID-19 pandemic. This study explored trajectories of anxiety and depression over the 20 weeks after lockdown was announced using data from England, and compared the growth trajectories by individual characteristics.

**Methods:** Data from 36,520 adults in the UCL COVID -19 Social Study (a panel study weighted to population proportions collecting data weekly during the COVID-19 pandemic) were analysed from 23/03/2020-09/08/2020. Latent growth models were fitted accounting for socio-demographic and health covariates.

**Findings:** The average depression score was 6.6 ± 6.0 in week 1 (range 0-27), while the average anxiety score was 5.7 ± 5.6 (range 0-21). Anxiety and depression levels both declined across the first 20 weeks following the introduction of lockdown in the England (b=-1.92, SE=0.26, p<.0001 & b=-2.52, SE=0.28, p<.0001). The fastest decreases were seen across the strict lockdown period, with symptoms plateauing as further lockdown easing measures were introduced. Being female or younger, having lower educational attainment, lower income or pre-existing mental health conditions, and living alone or with children were all risk factors for higher levels of anxiety and depression at the start of lockdown. Many of these inequalities in experiences were reduced as lockdown continued, but differences were still evident 20 weeks after the start of lockdown.

**Interpretation:** As countries face potential future lockdowns, these data suggest that the highest levels of depression and anxiety are in the early stages of lockdown but decline fairly rapidly as individuals adapt to circumstances. They also suggest the importance of supporting individuals in the lead-up to lockdown measures being brought in to try and reduce distress and highlight that emotionally vulnerable groups have remained at risk throughout lockdown and its aftermath.

**Funding:** This Covid-19 Social Study was funded by the Nuffield Foundation [WEL/FR-000022583], but the views expressed are those of the authors and not necessarily the Foundation. The study was also supported by the MARCH Mental Health Network funded by the Cross-Disciplinary Mental Health Network Plus initiative supported by UK Research and Innovation [ES/S002588/1], and by the Wellcome Trust [221400/Z/20/Z]. DF was funded by the Wellcome Trust [205407/Z/16/Z].

**Research in context:** *Evidence before this study:* We searched PubMed for articles published in English from 1 January 2020 to 14 September 2020 for studies published in English using the following keywords: (“COVID*” OR “coronavirus”) and (“anxiety” OR “depression” OR “mental health” OR “mental illness” OR “distress”). Studies using data from representative cohort studies revealed the substantial impact of the COVID-19 pandemic on levels of depression, anxiety and mental distress, showing increases in average scores of symptoms of psychological distress from before to during the pandemic as well as a rise in the proportion of people experiencing clinically significant levels of mental illness. But there was a gap in the evidence in understanding how mental health changed week by week over the course of the COVID-19 pandemic.

*Added value of this study:* This study extends previous findings that had shown that changes in mental health did occur by showing when these changes took place across first 20 weeks of restrictions in the UK and how the timing of changes corresponded to measures to ease lockdown. Further, this study highlights the inequalities in mental health across demographic and socio-economic groups and demonstrates how these inequalities changed over time.

*Implications of all the available evidence:* Overall, it is clear that mental health was adversely affected during the early months of the COVID-19 pandemic in the UK, especially in the early weeks of March 2020. However, once strict lockdown measures were brought in to control the virus, many people began to experience improvements in mental health. As countries face potential future lockdowns, these data suggest the importance of supporting individuals in the lead-up to lockdown measures being brought in to try and reduce distress but also suggest that individuals may be able to adapt relatively fast to the new psychological demands of life in lockdown. Many known risk factors for poorer mental health were apparent at the start of lockdown, but some groups experienced faster improvements in symptoms, thereby reducing the differences over time. Nevertheless, many inequalities in mental health experiences did remain and emotionally vulnerable groups have remained at risk throughout lockdown and its aftermath. These groups could benefit from more targeted mental health support as the pandemic continues.

## Introduction

There has been wide-spread concern for mental health during the COVID-19 pandemic been driven by the multiple different psychological challenges caused by the pandemic ^1^, and a call for urgent mental health research ^2^.. “Stay-at-home” and quarantine orders issued by governments led to the largest enforced isolation period in human history. Infections and deaths from the virus led to psychological stress and bereavement. Further, many individuals globally faced high levels of adversities, from challenges meeting basic needs (e,g. accessing food, water and safe accommodation), to financial adversities (including job losses, income cuts, and inability to pay bills) ^3^.

Data from representative cohort studies comparing data collected prior to the pandemic to data collected in the first few weeks of lockdowns internationally have shown increases in average scores of psychological distress and a rise in the proportion of people experiencing clinically significant levels of mental illness ^4,5^. These findings echo those from studies of previous epidemics such as SARS (severe acute respiratory syndrome), during which individuals who had to quarantine experienced increases in symptoms of depression and PTSD ^6,7 8 9^. However, what remains unclear is the trajectory of mental health across the course of the COVID-19 pandemic. The previous studies on SARS have suggested that mental health worsened *during* periods of quarantine or enforced isolation. However, a number of sources suggested that during COVID-19, mental health deteriorated *prior* to stay-at-home orders (“lockdown”) coming in ^10,11 12^. Given these findings, what remains to be understood is whether mental health continued to worsen as lockdown continued, or whether there were any patterns of stabilisation or improvement. Similarly, it is unknown as lockdown measures were eased, whether mental health improved or whether new stressors arose for individuals. These are vital questions as understanding the patterns of mental health across lockdowns could help mental health services and voluntary organisations to plan for future waves of the virus. Further, understanding how humans respond to periods of enforced isolation could enhance our understanding of the impact of social isolation on mental health.

Finally, an important question is whether certain individuals were more adversely affected during the COVID-19 pandemic. There are well-reported inequalities in mental health reported over the previous decade. Women, younger adults, individuals of lower educational attainment and socio-economic position, individuals from Black, Asian and Minority Ethnic (BAME) backgrounds, and individuals living alone are all more likely to experience higher levels of depression and anxiety ^13,14^. Data from previous epidemics have suggested that many of these factors have also been risk factors for worse mental health during periods of isolation ^15^, and data from COVID-19 has echoed this too ^16–21^. Similarly, there has been some indication that pre-existing psychiatric conditions are a risk factor for poorer outcomes^22^; also echoed by data during COVID-19 ^16,17^. However, previous research has focused on cross-sectional data or discrete time-points during the pandemic. The effect of such factors on trajectories of mental health remains unknown. Identifying these risk factors is important so as to be able to ascertain who is most in need of support both during the ongoing pandemic and in preparing for future pandemics.

Therefore, this study had two main aims: (i) to explore trajectories of anxiety and depressive symptoms over the “strict” lockdown period and as lockdown was eased, and (ii) to identify who was most at risk of poorer trajectories of mental health across this period.

## Methods

### Participants

Data were drawn from the UCL COVID-19 Social Study; a large panel study of the psychological and social experiences of over 70,000 adults (aged 18+) in the UK during the COVID-19 pandemic. The study commenced on 21^st^ March 2020 involving online weekly data collection from participants during the COVID-19 pandemic. Whilst not random, the study has a heterogenous sample that was recruited using three primary approaches. First, snowballing was used, including promoting the study through existing networks and mailing lists (including large databases of adults who had previously consented to be involved in health research across the UK such as UCL BioResource and HealthWise Wales, through Local Authorities and Mutual Aid groups across the UK, and via the UKRI Mental Health Research Networks), print and digital media coverage, and social media. Second, more targeted recruitment was undertaken through partnership work with recruitment companies (Find Out Now, SEO Works, FieldworkHub and Optimal Workshop) focusing on (i) individuals from a low-income background, (ii) individuals with no or few educational qualifications, and (iii) individuals who were unemployed. Third, the study was promoted via partnerships with third sector organisations within the UKRI MARCH Mental Health Research Network to vulnerable groups, including adults with pre-existing mental illness, older adults, and carers. Active recruitment was carried out for the first 8 weeks of the study. The study was approved by the UCL Research Ethics Committee [12467/005] and all participants gave informed consent. No participants received any payment to participate. Full detail on the recruitment, sampling, retention, and weighting of the sample is available in the Appendix and in the study User Guide (https://github.com/UCL-BSH/CSSUserGuide).

For these analyses, in order to examine trajectories of mental health in relation to specific measures relating to lockdown, we focused solely on participants who lived in England (n=59,348). Participants included in the analysis had at least three repeated measures between 23^rd^ March when lockdown started in the UK and 9^th^ August 2020 (20 weeks later). This provided us with data from 40,520 respondents who were followed over a maximum of 20 weeks since the beginning of the lockdown. Of these, 10% participants withheld data or preferred not to self-identify on demographic factors including gender and income so were excluded (the demographics of these participants are shown in Appendix p3), providing a final analytic sample size of 36,520.

### Measures

Depressive symptoms were measured using the Patient Health Questionnaire (PHQ-9); a standard 9-item instrument for diagnosing depression in primary care ^23^, with 4-point responses ranging from “not at all” to “nearly every day”. Scores of 0-4 suggest minimal depression, 5-9 mild depression, 10-14 moderate depression, 15-19 moderately severe depression, and 20-27 severe depression ^24^.

Anxiety was measured using the Generalised Anxiety Disorder assessment (GAD-7); a well-validated 7-item tool used to screen and diagnose generalised anxiety disorder in clinical practice and research ^25^ with 4-point responses ranging from “not at all” to “nearly every day”. Scores of 5-9 are thought to represent mild anxiety, 10-14 moderate anxiety, and 15 and above severe ^25^. Whilst the validated measures of both the PHQ-9 and GAD-7 ask respondents to focus on the past fortnight, as this study involved weekly reassessments, we asked participants to focus just on the past week.

We included socio-demographic variables as time-invariant covariates, including gender (men vs. women), age groups (18-29, 30-45, 46-59 and 60+), ethnicity (white vs. BAME), education (GCSE or below [equivalent to education to age 16], A levels or equivalent [equivalent to education to age 18], degree or above [further education after the age of 18]), low income (household income <£30,000 vs higher) and living arrangement (alone, living with others, but no children in the household, living with others, including children). We assessed diagnosed mental illness through participants “Do you have any of the following medical conditions”, with the responses including “clinically-diganosed depression”, “clinically-diagnosed anxiety” and “another clinically-diagnosed mental health problem”. Participants could select as many categories as applied and the responses were binarised into “diagnosed mental illness” or “no diagnosed mental illness”. Participants were also asked whether they had had COVID-19 (“yes, diagnosed and recovered/still ill”, “Not formally diagnosed but suspected”, or “Not that I know of / No”. However, only a very small percentage of the sample (0.02-0.88% each week) reported actually being formally diagnosed with a test due to limitations on testing in the early months of the pandemic so we did not include experience of COVID-19 in analyses. Further detail on the measures is available in the study User Guide (https://github.com/UCL-BSH/CSSUserGuide).

The timeline of key dates in lockdown restrictions in England is shown in dotted lines in Figure 1 and a description of the specific measures at key dates is shown in Appendix (p1).

**Figure 1.**
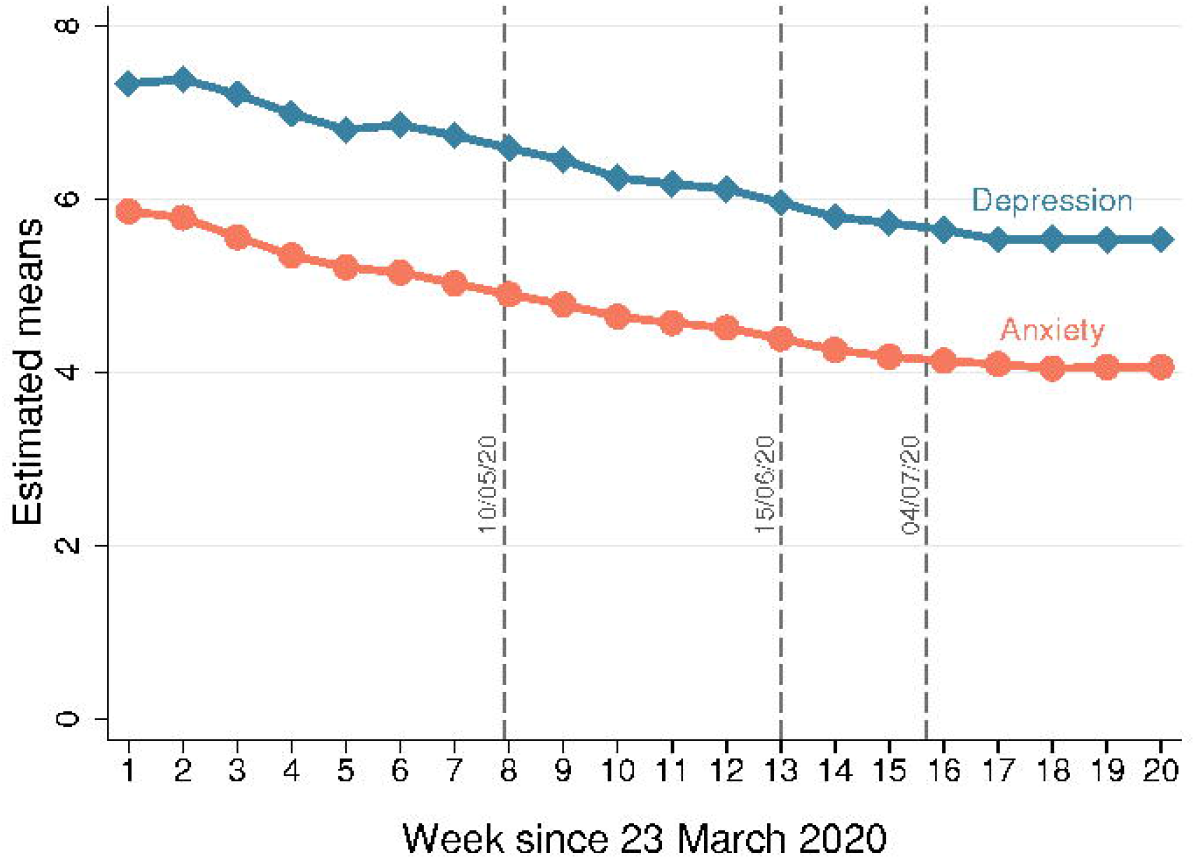
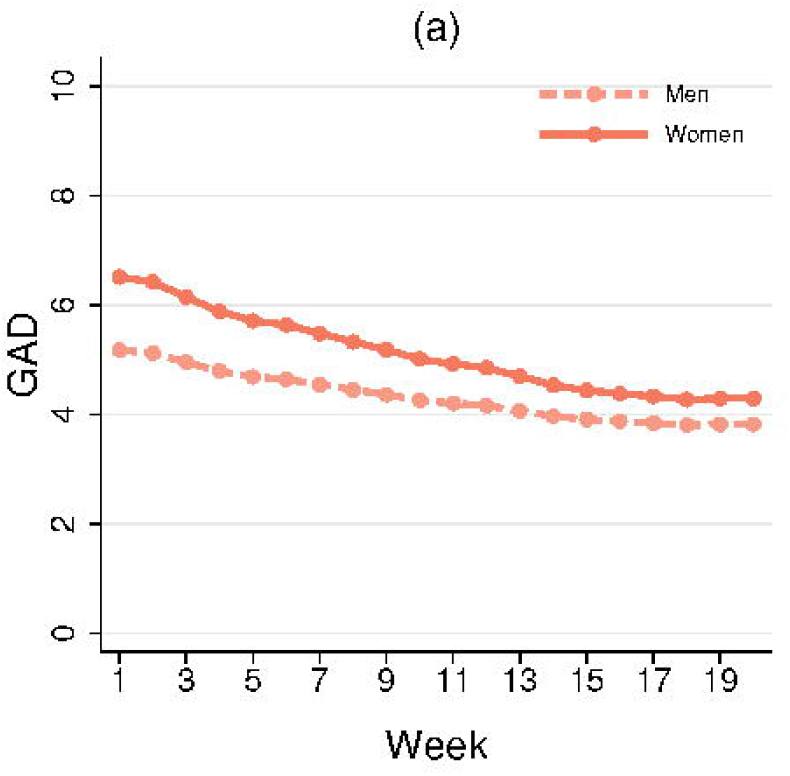
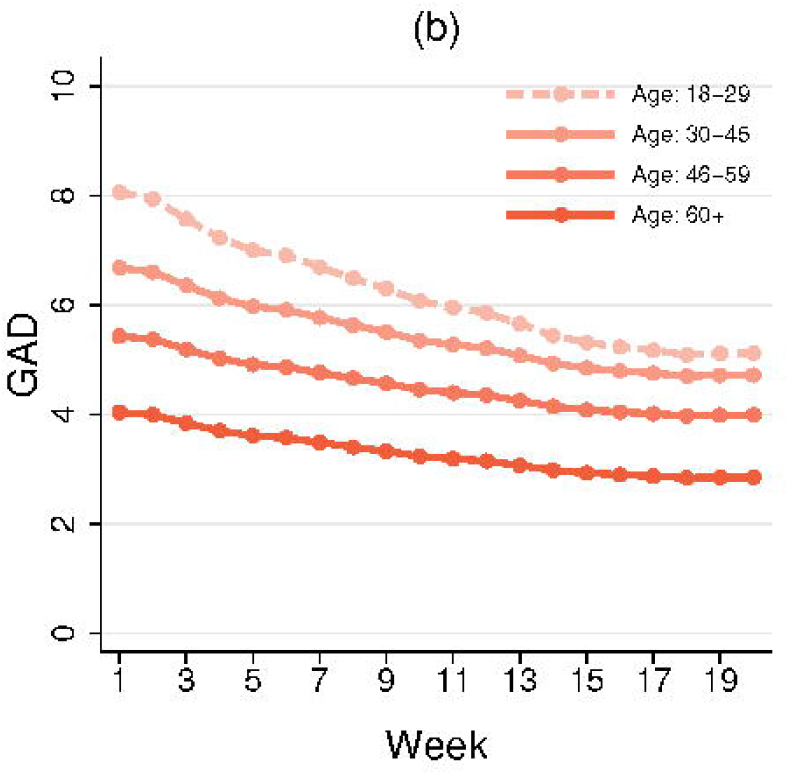
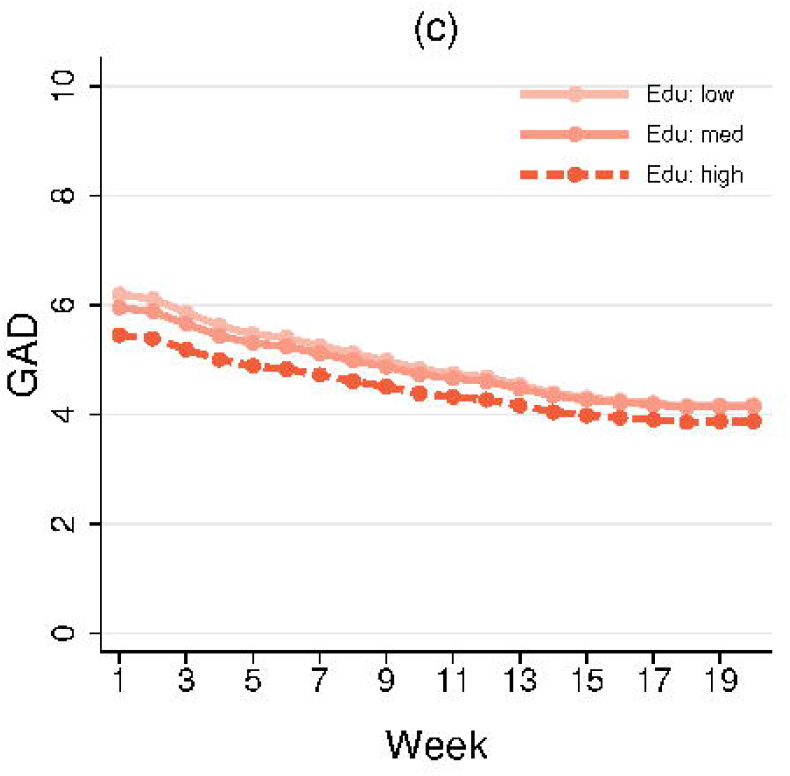
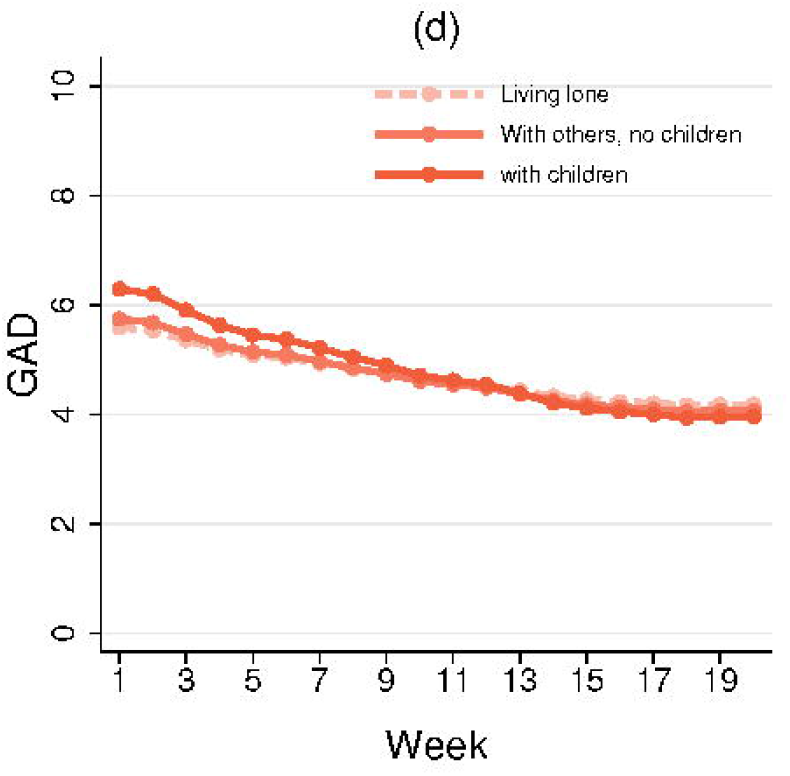
Predicted growth trajectories from the multi-process LGM model.

### Analysis

Data were analysed using latent growth modelling (LGM). We used unspecified LGM which allows the shape of growth trajectories to be determined by data by using free time scores. Given anxiety and depressive symptoms might be related, these two outcomes were modelled simultaneously (multi-process LGM). See Appendix p1 for an illustration of the model specification. The general equations were presented as below:

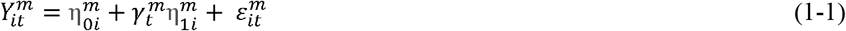

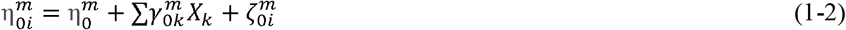

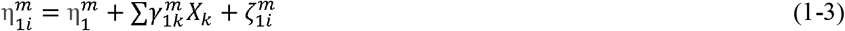

Equation 1-1 addressed intra-individual changes where 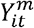, outcome *m* (depressive symptoms or anxiety) for the individual *i* at time *t*, was a function of the intercept 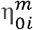 and slope 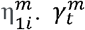 values were time scores 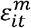 was the residual term. Equation 1-2, and 1-3 addressed inter-individual differences in the intercept and slope. 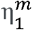 and 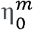 represented the population average intercept and slope for outcome *m*. X_k_ represented a vector of time-invariant variables that hypothetically influence the intercept and slope. 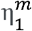 and 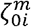 were parameter residuals. The predicted growth trajectories were based on the model estimates. Except for the grouping variable, all other variables were set to the sample means.

Our 36,520 participants provided a total of 436,522 observations (12 repeated measures per participant on average, ranging from 3 to 20). The distribution of follow-up weeks is shown in Appendix p3. To account for the non-random nature of the sample, the sample was weighted to the proportions of gender, age, ethnicity and education obtained from the Office for National Statistics ^26^. The correlation between outcome measures is shown in Appendix p3. Unweighted and weighted descriptive statistics by week are shown in Appendix p4. Descriptive analyses were carried out using Stata version 15. The LGM models were fitted in Mplus version 8.

### Patient and public involvement

The research questions in the COVID-19 Social Study built on patient and public involvement as part of the UKRI MARCH Mental Health Research Network, which highlighted priority research questions and measures for this study. Patients and the public were additionally involved in the recruitment of participants and the dissemination of findings.

## Results

In the raw data, 76% of participants (n=27,699) were women and there was an over-representation of people with a degree or above (70%; n=25,653) and an under-representation of people from BAME backgrounds (5%; n=1,839) (Table 1). After weighting, the sample reflected population proportions, with 51% women (n=18,643), 35% of participants with a degree or above (n=12,670), and 14% of BAME ethnicity (n=5,311). About 20% of the sample reported having a diagnosed mental illness (n=7,270).

In week 1, the average score on the PHQ-9 was 6.6 ± 6.0: 48% of the sample (n=8,228) had a score of 0-4 suggesting minimal depression, 27% had a score of 5-9 indicating mild depression (n=4,578), 13% had a score of 10-14 indicating moderate depression (n=2,218), 8% had a score of 15-19 indicating moderately severe depression (n=1,290), and 5% had a score of 20-27 indicating severe depression (n=776). The average score on the GAD-7 was 5.7 ± 5.6 in week 1: 53% of the sample (n=9,123) had a score of 0-4 suggesting minimal anxiety, 24% had a score of 5-9 indicating mild anxiety (n=4,111), 12% had a score of 10-14 indicating moderate anxiety (n=2,092), and 10% had a score of 15+ indicating severe anxiety (n=1,764) (Appendix p3). Amongst people with a pre-existing diagnosed mental illness, 54% (n=1,703) had a score ≥10 indicating at least moderate anxiety (average score 10.6 ± 5.8) and 61% (n=1,929) had a score ≥10 indicating moderate or severe depression (average score 12.3 ± 6.7). Amongst people without a pre-existing diagnosed mental illness, these figures were 16% (n=2,153) for anxiety (average score 4.6 ± 4.9) and 17% (n=2,356) for depression (average score 5.1 ± 5.0). More detail on symptoms by week is shown in Appendix p10.

Over the 20 weeks following the introduction of lockdown, there was a significant decrease in both anxiety and depressive symptoms both across strict lockdown and as lockdown was eased (Figure 1). The growth trajectories of anxiety and depressive symptoms showed a non-linear pattern (see Appendix p4). The slopes varied across different stages. For instance, there was a sharp decline in depressive symptoms and anxiety between weeks two and five during strict lockdown, but little change was observed between weeks 16 and 20 after substantial easing of lockdown had taken place. The growth trajectories of anxiety and depressive symptoms were positively associated with each other as evidenced by the significant covariance between the two intercepts (20.46, p<0.0001) and slopes (18.64, p<0.0001) (See Appendix p11).

At the start of lockdown being introduced, women, younger adults, people with lower levels of educational attainment, people from lower income households, and people with pre-existing mental health conditions reported higher levels of anxiety and depressive symptoms (Table 3). Individuals living with children had higher levels of anxiety (b=0.70, p=<.0001) but lower levels of depressive symptoms (b=-0.37, p=.029) compared to individuals living alone, whilst individuals living alone had higher levels of depressive symptoms but comparable levels of anxiety to people living with other adults (but no children). No evidence was found that ethnicity was associated with baseline mental health (anxiety: b=0.19, p=.43, depressive symptoms: b=0.45, p=.064).

Across the 20 weeks, women, younger adults, people with lower levels of educational attainment, and people living with children had faster improvements in symptoms of depressive symptoms and anxiety, narrowing some of the gaps in experiences present at the start of lockdown (Table 3 and Figures 2-3). People living alone experienced the same trajectories of depressive symptoms as people living with other adults (but no children) (living with others vs alone: b=0.04, p=.77), but their overall levels of depressive symptoms were consistently higher. There was no evidence that the rate of change was related to ethnicity, household income, or pre-existing mental health conditions. Inequalities by gender, age, education, income, and mental health were all still present at the end of the 20 weeks. For further graphs, see Appendix p2.

**Figure 2.**
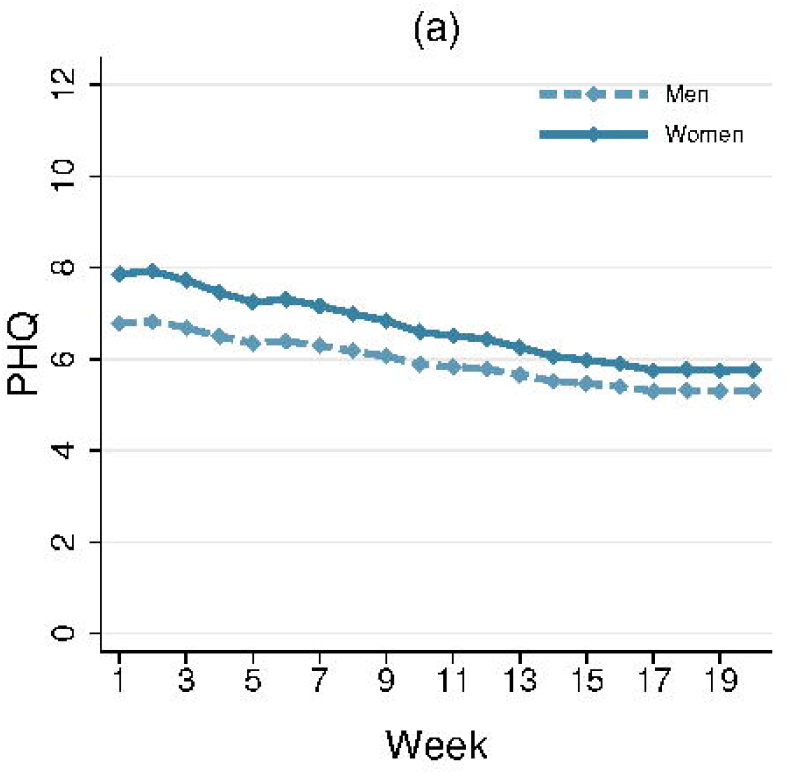
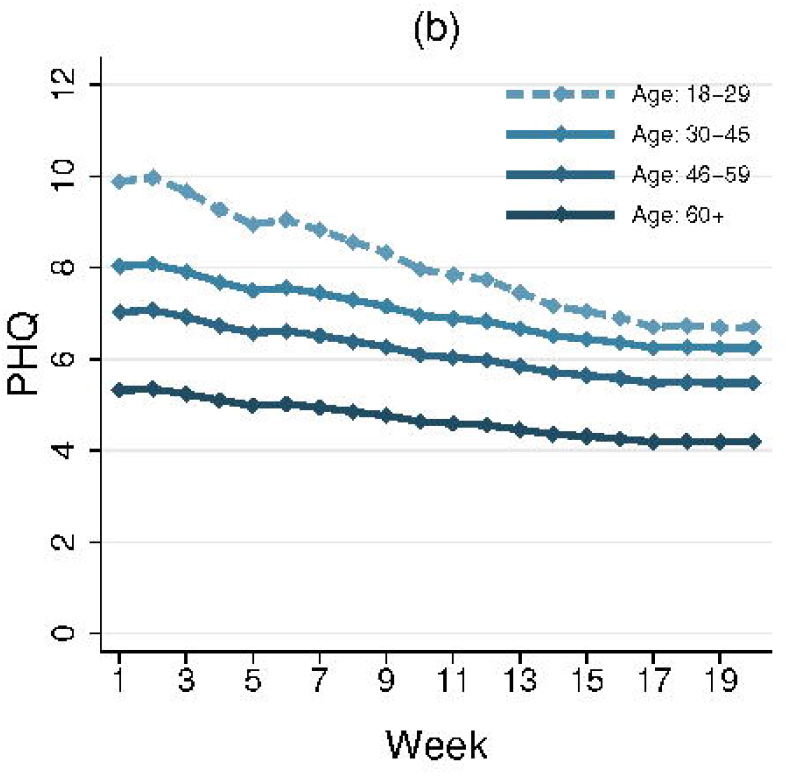
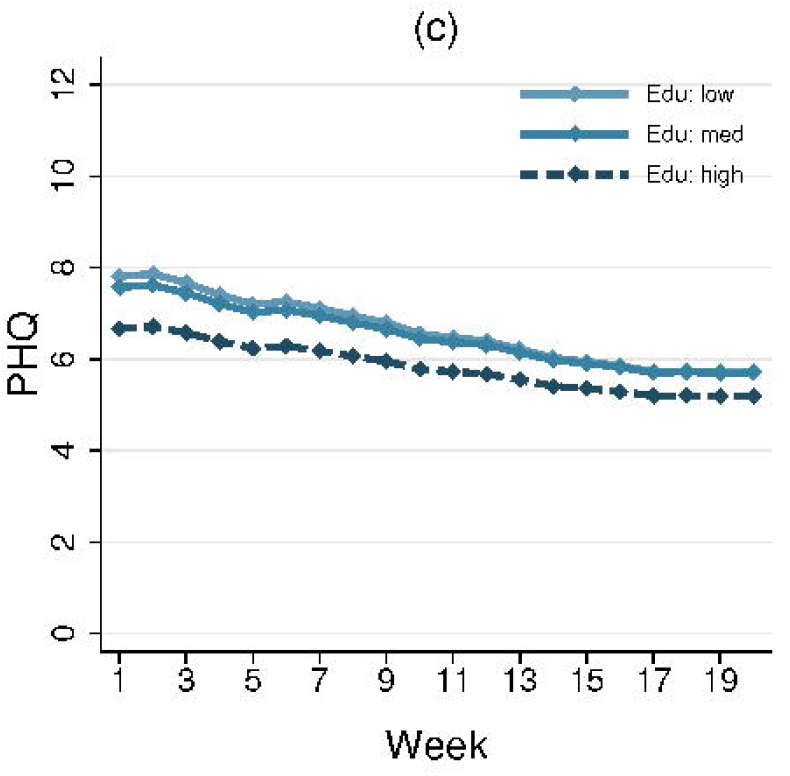
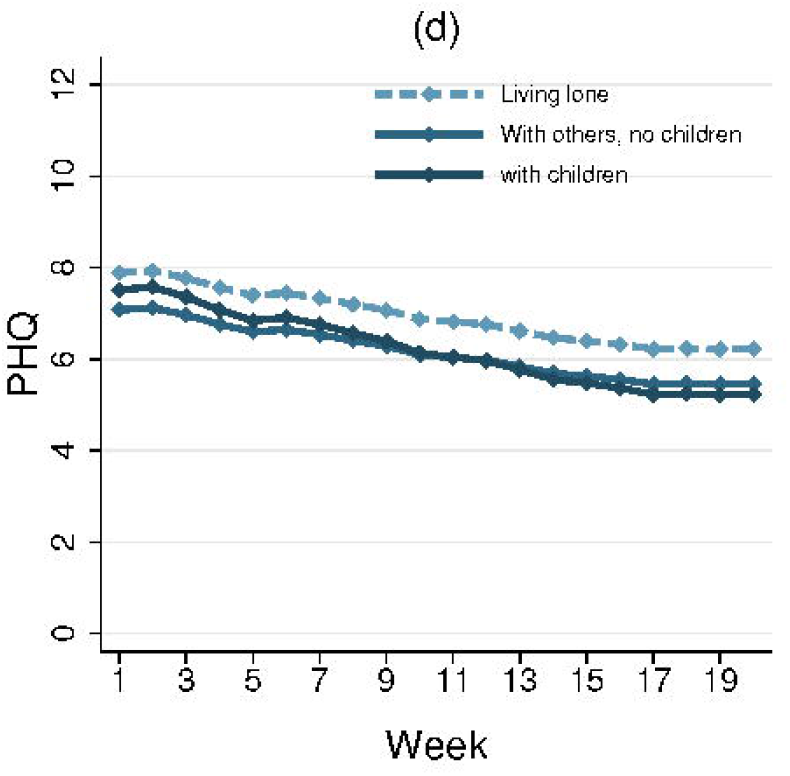
Predicted growth trajectories of anxiety by individual characteristics.

## Discussion

This study explored trajectories of anxiety and depression across the early stages of the lockdown response to the COVID-19 pandemic. Results show that anxiety and depressive symptoms both declined across the first 20 weeks following the introduction of lockdown in England. The fastest decreases were seen across the strict lockdown period, with symptoms plateauing as further lockdown easing measures were introduced. Being female or younger, having lower educational attainment, lower income, or pre-existing mental health conditions, and living alone or with children were all risk factors for higher levels of anxiety and depressive symptoms at the start of lockdown. Many of these inequalities in experiences were reduced as lockdown continued, but differences were still evident 20 weeks after the start of lockdown.

The study did not use a random population sample, and therefore our reported statistics are not presented as prevalence levels for anxiety or depressive symptoms. However, this study does give detailed time series data on trajectories of mental health during lockdown. The findings of improvements, in particular across the early weeks following the introduction of lockdown measures in England, is echoed by polling data showing improvements in many aspects of mood in repeated cross-sectional samples over the same period ^10,11^. The fact that levels of mental health did not continue to worsen even further in this period is slightly at odds with data from previous epidemics, in which mental health was found to worsen *during* (or as a result of) quarantine ^15^. However, there are several key differences between this pandemic and previous epidemics. First, for the majority of people in England during lockdown, some trips outside of the home were permissible, compared to quarantines studied in previous epidemics where movement has been even more restricted, albeit typically for much smaller numbers of people, which may have led to a harsher psychological experience. Second, there was substantial prior warning in England that a lockdown was likely to come in given patterns in countries across Europe, so individuals appear to have become psychologically affected prior to the lockdown announcement (and many individuals locked down voluntarily before lockdown officially commenced), meaning that much of the psychological toll was already being experienced before individuals were forced to isolate ^15^. Third, the proliferation of online and home-based leisure activities and the extensive use of virtual and digital communication during COVID-19 may have helped to ease the burden of lockdown itself, in contrast to previous epidemics where ‘fear of missing out’ was reported to be a challenge. Indeed, fear of missing (which is linked with depression, distraction, and somatic symptoms ^27^) out may also have been minimised due to the global nature of the pandemic, compared to the restricted nature of quarantines in previous epidemics. The improvements in mental health over this period also suggest a process of adaptation that bears similarities to literature on other types of isolation such as incarceration, where some studies have shown that depression levels can stabilise and even decrease on average month on month as new coping strategies emerge ^28^. It is further possible that measures to safeguard jobs and finances taken in the UK may have helped to reduce specific anxieties. The lockdown itself may also have reduced worries about individuals or their friends or families catching the virus, especially after the first two weeks of lockdown once individuals could be more confident they were outside of the incubation period.

This study also found there were substantial differences in experiences of mental health across the first 20 weeks following the introduction of lockdown amongst different groups. Previous studies during the COVID-19 pandemic have already highlighted that women and younger adults experienced higher levels of anxiety and depression ^4,17,18^, but our data show that these groups have also had faster improvements in their symptoms. This could indicate a more challenging psychological experience early on in lockdown (e.g. as many women balanced childcare and working from home) or a higher initial reactivity to events initially during lockdown amongst these groups. Adults living alone, on the other hand, experienced consistently worse levels of depressive symptoms, which could be related to higher levels of loneliness due to social restrictions ^29^. However, individuals living with children had higher levels of anxiety and depressive symptoms initially than individuals living with other adults, but a faster rate of improvement, potentially due to the growing public awareness of research suggesting that children were less at risk from the virus ^30^. Similarly, individuals with lower household income experienced consistently worse mental health, which has been proposed as related to higher experiences of adversities such as job losses, decreases in household income, and challenges paying bills ^3^. But differences in mental health at baseline are likely attributable to pre-existing social inequalities that have been exacerbated over the past decade ^31^. It is also notable that although we found that individuals with mental illness had higher levels of depressive symptoms and anxiety at the start of lockdown (echoing in studies from various countries of COVID-19 ^32^), they experienced the same trajectories over the subsequent weeks. This suggests that mental illness did not necessarily predispose individuals to greater levels of emotional reactivity. It is possible that prior experience of mental illness or social isolation caused by prior mental illness meant individuals had experience of some of the requirements of lockdown or of applying coping strategies in stressful situations. But lockdown may also have enabled unhealthy coping strategies, so this remains to be explored further. Further, future studies are recommended to look at specific type of psychiatric diagnosis in relation to mental health to identify if there are particular psychopathologies associated with poorer trajectories over this period. Our models suggested that ethnicity was not a risk factor for worse mental health. But it is important to note here that the models adjusted simultaneously for multiple different demographic factors including socio-economic position (SEP), which was related to poorer psychological experiences. Individuals from ethnic minority groups are disproportionately more likely to be from these lower SEP groups ^33^, so may still have been disproportionately affected. However, we use some binary categories in our analyses for ethnicity (as well as for other variables such as gender), but we recognise that such labels do not fully capture the experiences of different groups. We lacked sufficient statistical power to look at the experiences of specific ethnic groups and our analysis did not explore the experiences of individuals with non-binary gender identities, but we recognise the need for and encourage and support future research on these areas.

This study had several limitations. It is possible that the study did not adequately cover the full range of experiences. Therefore, this study does not claim to provide data on prevalence of mental illness during the pandemic: this question has been addressed through other representative studies. Further, as the study was internet based, participants without home access to internet were not represented. We also asked about symptoms of anxiety and depression over the past week rather than past fortnight as our reassessments were weekly. Whilst this change has precedence both in previous studies and clinical screening, it further underlines that the precise values on the scale cannot be taken as national averages for anxiety and depression during the pandemic. Additionally, we relied on participant self-report of diagnosed mental illness but were unable to confirm diagnoses, nor identify specifically the type of problem participants had previously been diagnosed with. As there was substantial overlap between participants reporting diagnoses of depression and diagnoses of anxiety, we also did not look at the unique contribution of each condition to trajectories of symptoms across lockdown, nor look at how specific symptoms (such as thoughts of death) might have been affected. As we asked about current diagnoses, we do not know how trajectories were affected by mental illness, specific psychological symptoms or sub-clinical symptomatology in the weeks or months preceding lockdown, and as participants entered the study continuously throughout the 20-week follow-up period reported here, it is possible that some diagnoses had arisen since lockdown began.. As this study involved repeated weekly assessment of mental health, regression to the mean is a further source of bias that warrants acknowledgement. However, if this is the case, we might have expected it to work both ways, with higher anxiety and depressive symptom scores declining and lower scores increasing. Yet all groups showed decreases across the study period. Further, a decrease in average scores due to non-random attrition is unlikely to have substantially biased results as all participants in the analysis provided at least three data points, so their trajectories were estimated even in the absence of complete data. Our study followed people across the period from spring to summer in England so the contribution of seasonality to findings remains to be explored further, although the results presented here suggest effects above and beyond usual fluctuations ^34^, and the validated measures of mental health used have been shown to have good test-retest reliability. Future studies could also consider how geographical factors including location within the UK, level of urbanisation, and area deprivation may have moderated psychological experiences during lockdown and whether experience of COVID-19 could have affected psychological response.

Overall, these findings suggest that the highest levels of depression and anxiety in England were in the early stages of lockdown but declined fairly rapidly following the introduction of lockdown, with improvements continuing as lockdown easing measures were introduced but plateauing after the first four months. Many known risk factors for poorer mental health were apparent at the start of lockdown, but some groups, including women, younger adults, and individuals with lower educational attainment experienced faster improvements in symptoms, thereby reducing the differences over time. Nevertheless, many inequalities in mental health experiences did remain and emotionally vulnerable groups have remained at risk throughout lockdown and its aftermath. As countries face potential future lockdowns, these data suggest the importance of supporting individuals in the lead-up to lockdown measures being brought in to try and reduce distress but also suggest that individuals may be able to adapt relatively fast to the new psychological demands of life in lockdown. But as inequalities in mental health have persisted, it is key to find ways of supporting vulnerable groups across this pandemic.

## Data Availability

Data will be made available following the end of the pandemic

## Declarations

### Ethics approval and consent to participate

Ethical approval for the COVID-19 Social Study was granted by the UCL Ethics Committee. All participants provided fully informed consent. The study is GDPR compliant.

### Data availability

Anonymous data will be made publicly available following the end of the pandemic.

### Author contributions

DF, AS and FB conceived and designed the study. FB analysed the data and DF wrote the first draft. All authors provided critical revisions. All authors read and approved the submitted manuscript.

## References

1 Holmes EA, O’Connor RC, Perry VH, et al. Multidisciplinary research priorities for the COVID-19 pandemic: a call for action for mental health science. The Lancet Psychiatry 2020; 0. DOI:10.1016/S2215-0366(20)30168-1.

2 Mahase E. Covid-19: Mental health consequences of pandemic need urgent research, paper advises. BMJ 2020; 369. DOI:10.1136/bmj.m1515.

3 Wright L, Steptoe A, Fancourt D. Are we all in this together? Longitudinal assessment of cumulative adversities by socio-economic position in the first 3 weeks of lockdown in the UK. Journal of Epidemiology & Community Health 2020; 74: 683–8.

4 Pierce M, Hope H, Ford T, et al. Mental health before and during the COVID-19 pandemic: a longitudinal probability sample survey of the UK population. The Lancet Psychiatry 2020; 0. DOI:10.1016/S2215-0366(20)30308-4.

5 Shanahan L, Steinhoff A, Bechtiger L, et al. Emotional distress in young adults during the COVID-19 pandemic: evidence of risk and resilience from a longitudinal cohort study. Psychological Medicine 2020; : 1–10.

6 Hawryluck L, Gold WL, Robinson S, Pogorski S, Galea S, Styra R. SARS Control and Psychological Effects of Quarantine, Toronto, Canada. Emerg Infect Dis 2004; 10: 1206–12.

7 Reynolds DL, Garay JR, Deamond SL, Moran MK, Gold W, Styra R. Understanding, compliance and psychological impact of the SARS quarantine experience. Epidemiology & Infection 2008; 136: 997–1007.

8 Mihashi M, Otsubo Y, Yinjuan X, Nagatomi K, Hoshiko M, Ishitake T. Predictive factors of psychological disorder development during recovery following SARS outbreak. Health Psychology 2009; 28: 91–100.

9 Liu X, Kakade M, Fuller CJ, et al. Depression after exposure to stressful events: lessons learned from the severe acute respiratory syndrome epidemic. Comprehensive Psychiatry 2012; 53: 15–23.

10 YouGov. Britain’s mood, measured weekly. https://yougov.co.uk/topics/science/trackers/britains-mood-measured-weekly (accessed April 20, 2020).

11 Layard R, Clark A, De Neve J-E, et al. When to release the lockdown: A wellbeing framework for analysing costs and benefits. Centre for Economic Performance, London School of Economics, 2020.

12 Millions of UK adults have felt panicked, afraid and unprepared as a result of the coronavirus pandemic - new poll data reveal impact on mental health. Mental Health Foundation. 2020; published online March https://www.mentalhealth.org.uk/news/millions-uk-adults-have-felt-panicked-afraid-and-unprepared-result-coronavirus-pandemic-new (accessed April 24, 2020).

13 Ansseau M, Fischler B, Dierick M, Albert A, Leyman S, Mignon A. Socioeconomic correlates of generalized anxiety disorder and major depression in primary care: the GADIS II study (Generalized Anxiety and Depression Impact Survey II). Depression and Anxiety 2008; 25: 506–513.

14 Hölzel L, Härter M, Reese C, Kriston L. Risk factors for chronic depression—a systematic review. Journal of affective disorders 2011; 129: 1–13.

15 Brooks SK, Webster RK, Smith LE, et al. The psychological impact of quarantine and how to reduce it: rapid review of the evidence. The Lancet 2020; 395: 912–20.

16 Kwong ASF, Pearson RM, Adams MJ, et al. Mental health during the COVID-19 pandemic in two longitudinal UK population cohorts. medRxiv 2020; : 2020.06.16.20133116.

17 Solomou I, Constantinidou F. Prevalence and Predictors of Anxiety and Depression Symptoms during the COVID-19 Pandemic and Compliance with Precautionary Measures: Age and Sex Matter. International Journal of Environmental Research and Public Health 2020; 17: 4924.

18 Pieh C, Budimir S, Probst T. The effect of age, gender, income, work, and physical activity on mental health during coronavirus disease (COVID-19) lockdown in Austria. Journal of Psychosomatic Research 2020; 136: 110186.

19 Pierce M, Hope H, Ford T, et al. Mental Health Before and During the COVID-19 Pandemic: A Longitudinal Probability Sample Survey of the UK Population. Rochester, NY: Social Science Research Network, 2020 https://papers.ssrn.com/abstract=3624264 (accessed July 9, 2020).

20 Wang C, Pan R, Wan X, et al. Immediate Psychological Responses and Associated Factors during the Initial Stage of the 2019 Coronavirus Disease (COVID-19) Epidemic among the General Population in China. International Journal of Environmental Research and Public Health 2020; 17: 1729.

21 Ausín B, González-Sanguino C, Castellanos MÁ, Muñoz M. Gender-related differences in the psychological impact of confinement as a consequence of COVID-19 in Spain. Journal of Gender Studies 2020; 0: 1–10.

22 Jeong H, Yim HW, Song Y-J, et al. Mental health status of people isolated due to Middle East Respiratory Syndrome. Epidemiol Health 2016; 38. DOI:10.4178/epih.e2016048.

23 Löwe B, Kroenke K, Herzog W, Gräfe K. Measuring depression outcome with a brief self-report instrument: sensitivity to change of the Patient Health Questionnaire (PHQ-9). Journal of Affective Disorders 2004; 81: 61–6.

24 Kroenke K, Spitzer RL. The PHQ-9: A New Depression Diagnostic and Severity Measure. Psychiatr Ann 2002; 32: 509–15.

25 Spitzer RL, Kroenke K, Williams JB, Löwe B. A brief measure for assessing generalized anxiety disorder: the GAD-7. Archives of internal medicine 2006; 166: 1092–1097.

26 Population estimates for the UK, England and Wales, Scotland and Northern Ireland - Office for National Statistics. https://www.ons.gov.uk/peoplepopulationandcommunity/populationandmigration/populationestimates/bulletins/annualmidyearpopulationestimates/mid2018 (accessed May 13, 2020).

27 Baker ZG, Krieger H, LeRoy AS. Fear of missing out: Relationships with depression, mindfulness, and physical symptoms. Translational Issues in Psychological Science 2016; 2: 275.

28 Porter LC, DeMarco LM. Beyond the dichotomy: Incarceration dosage and mental health*. Criminology 2019; 57: 136–56.

29 Bu F, Steptoe A, Fancourt D. Who is lonely in lockdown? Cross-cohort analyses of predictors of loneliness before and during the COVID-19 pandemic. Public Health 2020; 186: 31–4.

30 Guan W, Ni Z, Hu Y, et al. Clinical Characteristics of Coronavirus Disease 2019 in China. New England Journal of Medicine 2020; published online Feb 28. DOI:10.1056/NEJMoa2002032.

31 Marmot M. Health equity in England: the Marmot review 10 years on. Bmj 2020; 368.

32 Newby JM, O’Moore K, Tang S, Christensen H, Faasse K. Acute mental health responses during the COVID-19 pandemic in Australia. PLOS ONE 2020; 15: e0236562.

33 Phillips C. Institutional Racism and Ethnic Inequalities: An Expanded Multilevel Framework. Journal of Social Policy 2011; 40: 173–92.

34 Harmatz MG, Well AD, Overtree CE, Kawamura KY, Rosal M, Ockene IS. Seasonal Variation of Depression and Other Moods: A Longitudinal Approach. J Biol Rhythms 2000; 15: 344–50.

