## Supplementary material for "Trajectories of anxiety and depressive symptoms during enforced isolation due to COVID-19: longitudinal analyses of 36,520 adults in England": Tables

Table 1: Descriptive statistics of the explanatory variables for participants at their baseline assessment

|  | Raw data | | Weighted data | |
| --- | --- | --- | --- | --- |
|  | Percent | N | Percent | N |
| Gender, women |  |  |  |  |
| Women | 75.8% | 27699 | 51.0% | 18643 |
| Men | 24.2% | 8821 | 49.0% | 17877 |
| Age |  |  |  |  |
| 18-29 | 7.5% | 2730 | 19.5% | 7130 |
| 30-45 | 29.2% | 10649 | 26.4% | 9643 |
| 46-59 | 33.0% | 12048 | 24.1% | 8805 |
| 60+ | 30.4% | 11093 | 30.0% | 10943 |
| Ethnicity, BAME | 5.0% | 1839 | 13.5% | 5311 |
| Education |  |  |  |  |
| GCSE or below | 13.0% | 4731 | 32.4% | 11848 |
| A-levels or equivalent | 16.8% | 6136 | 32.9% | 12003 |
| Degree or above | 70.2% | 25653 | 34.7% | 12670 |
| Household income <30k (VS. ≥30k) | 36.7% | 13417 | 46.1% | 16847 |
| Living status |  |  |  |  |
| Alone | 19.7% | 7195 | 18.3% | 6684 |
| With others (not children) | 53.2% | 19411 | 56.1% | 20483 |
| With others (including children) | 27.1% | 9914 | 25.6% | 9352 |
| Diagnosed mental illness | 18.3% | 6679 | 19.9% | 7270 |

Table 2: Levels of anxiety and depression in the sample in week 1 (weighted)

|  | **Anxiety** | | **Depression** | |
| --- | --- | --- | --- | --- |
|  | Mean (SD) | % ≥10 (N) | Mean (SD) | % ≥10 (N) |
| **Total sample** | 5.7 (5.6) | 22.6% (3856) | 6.6 (6.0) | 25.1% (4285) |
| **Gender** |  |  |  |  |
| Men | 4.3 (3.5) | 14.1% (1240) | 5.4 (4.0) | 18.6% (1635) |
| Women | 7.2 (7.3) | 31.5% (2616) | 7.5 (7.7) | 31.9% (2649) |
| **Age** |  |  |  |  |
| 18-29 | 7.6 (3.8) | 34.4% (1054) | 8.3 (4.0) | 32.7% (1003) |
| 30-45 | 7.2 (6.2) | 30.4% (1421) | 7.5 (6.5) | 30.9% (1448) |
| 46-59 | 5.8 (6.4) | 21.4% (875) | 6.9 (7.3) | 27.8% (1138) |
| 60+ | 3.2 (4.1) | 9.6% (505) | 4.2 (4.7) | 13.3% (697) |
| **Ethnicity** |  |  |  |  |
| White | 5.7 (5.9) | 22.7% (3337) | 6.4 (6.4) | 25.1% (3697) |
| BAME | 5.7 (3.2) | 21.8% (519) | 6.9 (3.3) | 24.7% (588) |
| **Education** |  |  |  |  |
| GCSE or below | 5.4 (3.7) | 21.5% (1187) | 6.4 (4.0) | 26.0% (1439) |
| A-levels or equivalent | 6.0 (4.2) | 25.2% (1414) | 7.0 (4.7) | 28.1% (1579) |
| Degree or above | 5.6 (7.3) | 21.1% (1255) | 6.0 (7.5) | 21.3% (1267) |
| **Household income** |  |  |  |  |
| ≥30k | 5.5 (5.8) | 20.2% (1838) | 5.7 (5.9) | 19.7% (1788) |
| <30k | 5.9 (5.2) | 25.2% (2018) | 7.3 (5.9) | 31.2% (2496) |
| **Living status** |  |  |  |  |
| Alone | 5.1 (5.4) | 20.5% (648) | 7.1 (6.6) | 29.3% (925) |
| With others (not children) | 5.3 (5.3) | 20.6% (1998) | 5.9 (5.6) | 22.0% (2134) |
| With others (including children) | 6.9 (6.1) | 28.5% (1210) | 7.3 (6.4) | 28.8% (1226) |
| **Diagnosed mental illness** |  |  |  |  |
| No | 4.6 (4.9) | 15.5% (2153) | 5.1 (5.0) | 16.9% (2356) |
| Yes | 10.6 (5.8) | 53.7% (1703) | 12.3 (6.7) | 60.9% (1929) |

NB not all participants started the study in week 1, so this does not represent the full N

Table 3: Estimated effects of the covariates on the intercepts and slopes from the conditional multi-process LGM model

|  | Anxiety | | | Depressive symptoms | | |
| --- | --- | --- | --- | --- | --- | --- |
|  | b | se | p | b | se | p |
| **Predictors of the intercept** |  |  |  |  |  |  |
| Women (VS. men) | **1.34** | **0.13** | **<0.0001** | **1.08** | **0.13** | **<0.0001** |
| Age: 30-45 (VS. 18-29) | **-1.38** | **0.22** | **<0.0001** | **-1.86** | **0.23** | **<0.0001** |
| Age: 46-59 (VS. 18-29) | **-2.63** | **0.23** | **<0.0001** | **-2.86** | **0.24** | **<0.0001** |
| Age: 60+ (VS. 18-29) | **-4.02** | **0.23** | **<0.0001** | **-4.57** | **0.25** | **<0.0001** |
| BAME (VS. white) | 0.19 | 0.24 | 0.434 | 0.45 | 0.24 | 0.064 |
| Education: low (VS. high) | **0.75** | **0.14** | **<0.0001** | **1.13** | **0.16** | **<0.0001** |
| Education: medium (VS. high) | **0.50** | **0.13** | **<0.0001** | **0.90** | **0.13** | **<0.0001** |
| Household income <30k (VS. ≥30k) | **0.65** | **0.12** | **<0.0001** | **1.09** | **0.13** | **<0.0001** |
| Living with others, no children (VS. alone) | 0.16 | 0.13 | 0.212 | **-0.81** | **0.14** | **<0.0001** |
| Living with others, incl. children (VS. alone) | **0.70** | **0.15** | **<0.0001** | **-0.37** | **0.17** | **0.029** |
| Mental health diagnosis (VS. none) | **5.18** | **0.18** | **<0.0001** | **5.83** | **0.18** | **<0.0001** |
| **Predictors of the Slope** |  |  |  |  |  |  |
| Women (VS. men) | **-0.86** | **0.13** | **<0.0001** | **-0.63** | **0.14** | **<0.0001** |
| Age: 30-45 (VS. 18-29) | **0.98** | **0.28** | **<0.0001** | **1.40** | **0.29** | **<0.0001** |
| Age: 46-59 (VS. 18-29) | **1.50** | **0.27** | **<0.0001** | **1.63** | **0.29** | **<0.0001** |
| Age: 60+ (VS. 18-29) | **1.75** | **0.27** | **<0.0001** | **2.06** | **0.28** | **<0.0001** |
| BAME (VS. white) | 0.18 | 0.27 | 0.507 | -0.01 | 0.29 | 0.980 |
| Education: low (VS. high) | **-0.45** | **0.15** | **0.003** | **-0.60** | **0.16** | **<0.0001** |
| Education: medium (VS. high) | **-0.23** | **0.14** | **0.112** | **-0.39** | **0.15** | **0.011** |
| Household income <30k (VS. ≥30k) | 0.17 | 0.13 | 0.186 | 0.13 | 0.14 | 0.342 |
| Living with others, no child (VS. alone) | **-0.27** | **0.13** | **0.036** | 0.04 | 0.14 | 0.773 |
| Living with others, incl. children (VS. alone) | **-0.93** | **0.18** | **<0.0001** | **-0.63** | **0.19** | **0.0001** |
| Mental health diagnosis (VS. none) | -0.32 | 0.20 | 0.117 | 0.28 | 0.22 | 0.193 |
