## Supplementary Material for "Trajectories of anxiety and depressive symptoms during enforced isolation due to COVID-19: longitudinal analyses of 36,520 adults in England"

Appendix

### Lockdown timeline

From weeks 1-8 of lockdown, individuals could only leave their home for exercise once a day, purchasing essentials, and essential work. On 10/05/20, strict lockdown was first eased, with unlimited outdoor exercise permitted, more movement around the country permitted, and some returns to work allowed. On 15/06/20, non-essential retail was reopened, with individuals allowed to go shopping again, secondary schools were opened for pupils in some years, and individuals living alone could form a ‘support bubble’ with another household. On 04/07/20, further public amenities reopened such as places of worship, museums, hairdressers, holiday accommodation, and cafes, pubs and restaurants, and individuals from two households could meet socially indoors.

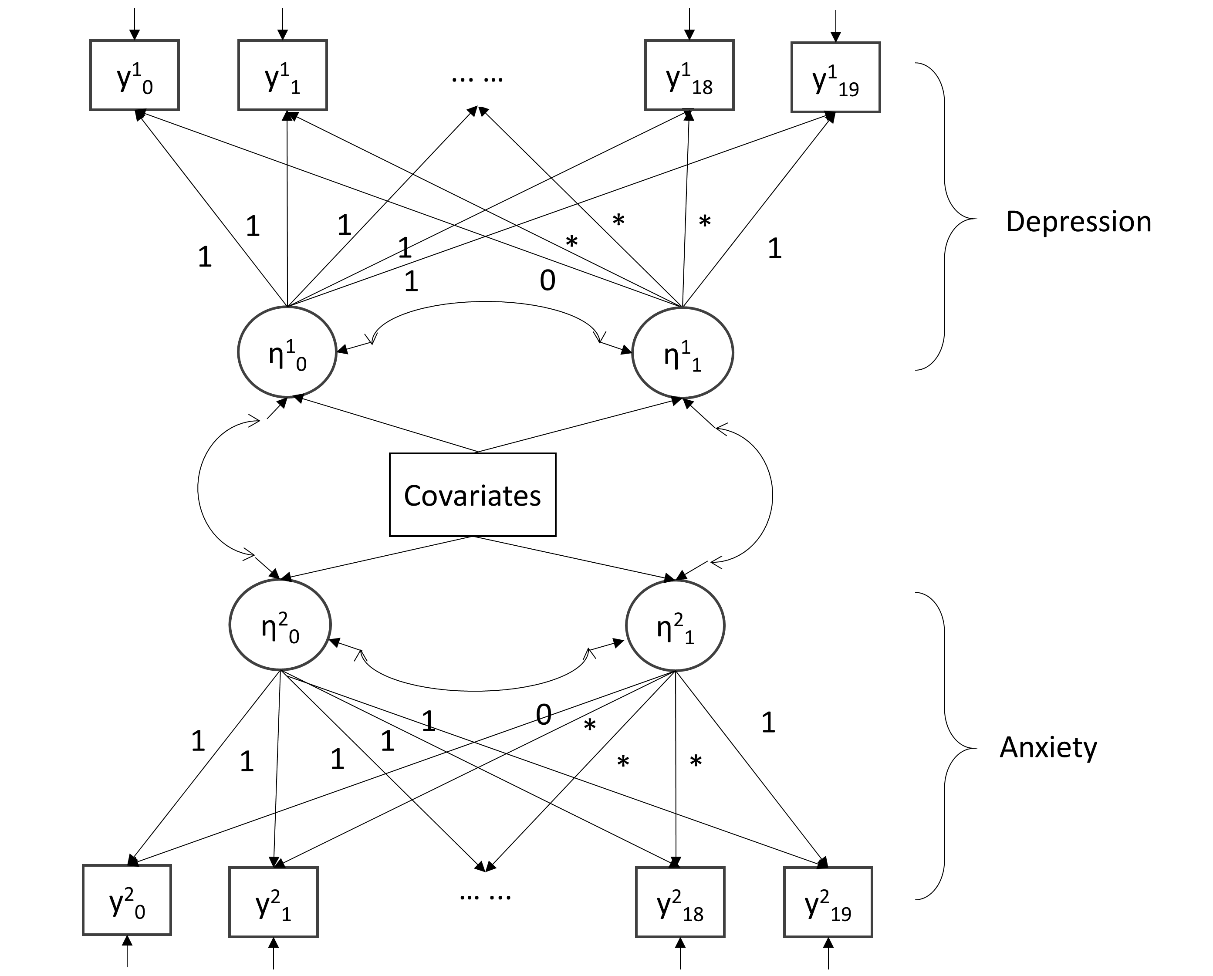

Figure S1. Conditional multi-process LGM

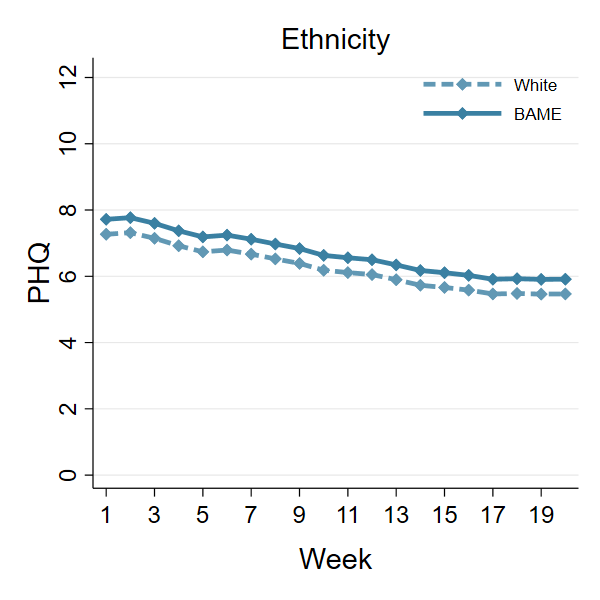

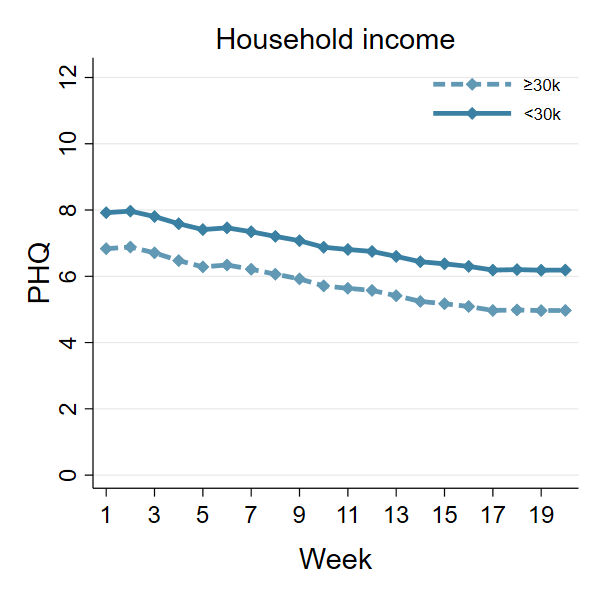

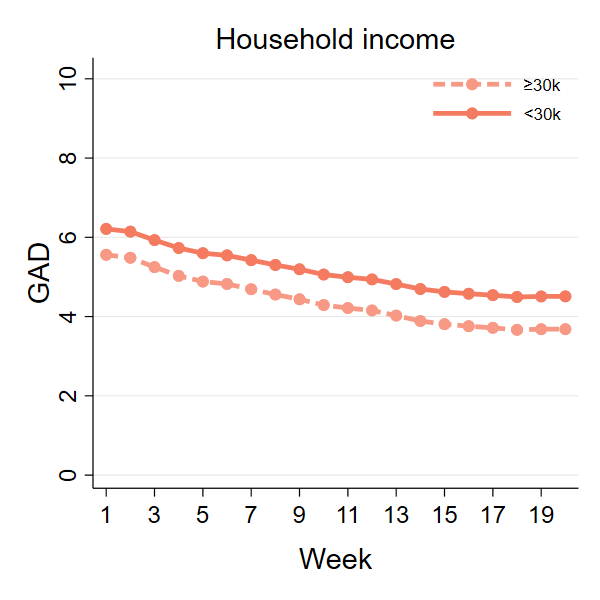

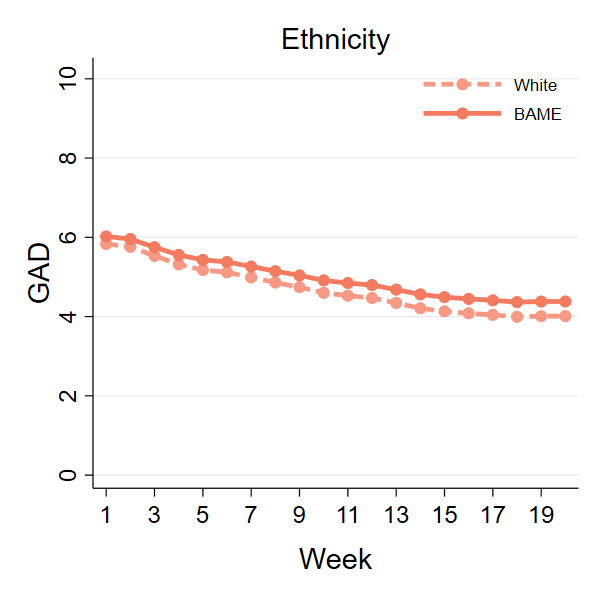

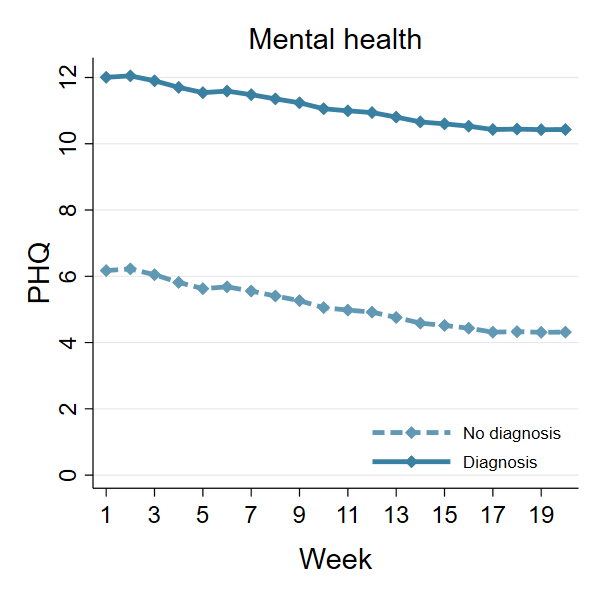

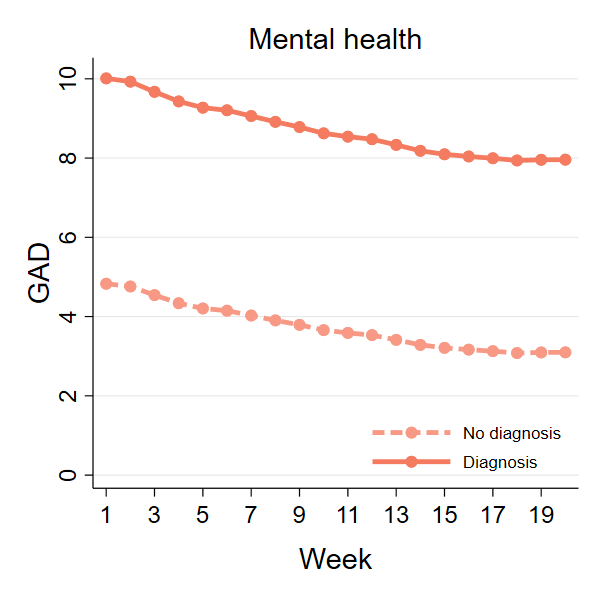

Figure S2: Predicted growth trajectories of anxiety by further individual characteristics

Table S1 Characteristics of participants who were excluded from the analysis due to missing data.

|  | Excluded participants (N=4000) | |
| --- | --- | --- |
|  | % | N |
| Gender, women | 80.8% | 3,835 |
| Age |  |  |
| 18-29 | 10.5% | 4,000 |
| 30-45 | 18.9% | 4,000 |
| 46-59 | 32.9% | 4,000 |
| 60+ | 37.8% | 4,000 |
| Ethnicity, BAME | 5.7% | 3,867 |
| Education |  |  |
| GCSE or below | 18.3% | 4,000 |
| A-levels or equivalent | 21.3% | 4,000 |
| Degree or above | 60.4% | 4,000 |
| Household income <30k | 51.2% | 213 |
| Living status |  |  |
| Alone | 14.0% | 4,000 |
| With others (not children) | 64.7% | 4,000 |
| With others (including children) | 21.3% | 4,000 |
| Diagnosed mental illness | 18.1% | 4,000 |

Table S2. The distribution of number of follow-up weeks

| Number of weeks | N | Percent |
| --- | --- | --- |
| 3 | 3,608 | 9.88 |
| 4 | 2,634 | 7.21 |
| 5 | 2,346 | 6.42 |
| 6 | 1,638 | 4.49 |
| 7 | 1,381 | 3.78 |
| 8 | 1,126 | 3.08 |
| 9 | 1,003 | 2.75 |
| 10 | 1,055 | 2.89 |
| 11 | 1,040 | 2.85 |
| 12 | 1,393 | 3.81 |
| 13 | 1,696 | 4.64 |
| 14 | 1,853 | 5.07 |
| 15 | 1,516 | 4.15 |
| 16 | 2,050 | 5.61 |
| 17 | 3,162 | 8.66 |
| 18 | 3,386 | 9.27 |
| 19 | 3,695 | 10.12 |
| 20 | 1,938 | 5.31 |

Table S3 Correlation between PHQ and GAD scores in by week

| Week | Correlation between GAD and PHQ |
| --- | --- |
| 1 | 0.76 |
| 2 | 0.78 |
| 3 | 0.79 |
| 4 | 0.81 |
| 5 | 0.81 |
| 6 | 0.83 |
| 7 | 0.83 |
| 8 | 0.83 |
| 9 | 0.83 |
| 10 | 0.83 |
| 11 | 0.83 |
| 12 | 0.84 |
| 13 | 0.83 |
| 14 | 0.84 |
| 15 | 0.84 |
| 16 | 0.84 |
| 17 | 0.84 |
| 18 | 0.84 |
| 19 | 0.84 |
| 20 | 0.85 |

Table S4 Unweighted and weighted descriptive statistics by week

|  | Week 1 | | | Week2 | | |
| --- | --- | --- | --- | --- | --- | --- |
|  | Raw | | Weighted | Raw | | Weighted |
|  | N | % | % | N | % | % |
| Gender, women | 12491 | 74.4% | 48.7% | 14569 | 75.3% | 50.2% |
| Age |  |  |  |  |  |  |
| 18-29 | 1269 | 7.6% | 17.9% | 1517 | 7.8% | 18.6% |
| 30-45 | 5161 | 30.7% | 27.4% | 6003 | 31.0% | 27.9% |
| 46-59 | 5328 | 31.7% | 23.9% | 6113 | 31.6% | 23.5% |
| 60+ | 5036 | 30.0% | 30.8% | 5717 | 29.5% | 30.0% |
| Ethnicity, BAME | 781 | 4.7% | 13.9% | 977 | 5.0% | 14.9% |
| Education |  |  |  |  |  |  |
| GCSE or below | 2211 | 13.2% | 32.4% | 2481 | 12.8% | 32.0% |
| A-levels or equivalent | 2869 | 17.1% | 32.8% | 3206 | 16.6% | 32.2% |
| Degree or above | 11714 | 69.8% | 34.8% | 13663 | 70.6% | 35.9% |
| Household income <30k | 6196 | 36.9% | 46.8% | 7000 | 36.2% | 46.0% |
| Living status |  |  |  |  |  |  |
| Alone | 3239 | 19.3% | 18.5% | 3702 | 19.1% | 18.1% |
| With others (not children) | 9022 | 53.7% | 56.7% | 10415 | 53.8% | 56.6% |
| With others (including children) | 4533 | 27.0% | 24.9% | 5233 | 27.0% | 25.3% |
| Diagnosed mental illness | 2968 | 17.7% | 18.5% | 3348 | 17.3% | 18.5% |

|  | Week 3 | | | Week 4 | | |
| --- | --- | --- | --- | --- | --- | --- |
|  | Raw | | Weighted | Raw | | Weighted |
|  | N | % | % | N | % | % |
| Gender, women | 19425 | 74.7% | 48.9% | 19843 | 75.0% | 49.3% |
| Age |  |  |  |  |  |  |
| 18-29 | 1856 | 7.1% | 17.4% | 1866 | 7.1% | 17.3% |
| 30-45 | 7754 | 29.8% | 27.0% | 7799 | 29.5% | 26.7% |
| 46-59 | 8497 | 32.7% | 24.3% | 8639 | 32.7% | 24.2% |
| 60+ | 7913 | 30.4% | 31.3% | 8148 | 30.8% | 31.8% |
| Ethnicity, BAME | 1201 | 4.6% | 13.1% | 1219 | 4.6% | 13.2% |
| Education |  |  |  |  |  |  |
| GCSE or below | 3541 | 13.6% | 33.7% | 3535 | 13.4% | 33.6% |
| A-levels or equivalent | 4526 | 17.4% | 32.8% | 4537 | 17.2% | 32.2% |
| Degree or above | 17953 | 69.0% | 33.5% | 18380 | 69.5% | 34.1% |
| Household income <30k | 9698 | 37.3% | 46.9% | 9755 | 36.9% | 46.4% |
| Living status |  |  |  |  |  |  |
| Alone | 5033 | 19.3% | 18.6% | 5218 | 19.7% | 19.0% |
| With others (not children) | 13886 | 53.4% | 55.9% | 14111 | 53.3% | 56.1% |
| With others (including children) | 7101 | 27.3% | 25.6% | 7123 | 26.9% | 25.0% |
| Diagnosed mental illness | 4640 | 17.8% | 18.7% | 4683 | 17.7% | 18.6% |

|  | Week 5 | | | Week 6 | | |
| --- | --- | --- | --- | --- | --- | --- |
|  | Raw | | Weighted | Raw | | Weighted |
|  | N | % | % | N | % | % |
| Gender, women | 19322 | 74.9% | 49.5% | 18716 | 75.4% | 50.4% |
| Age |  |  |  |  |  |  |
| 18-29 | 1830 | 7.1% | 17.9% | 1675 | 6.7% | 17.6% |
| 30-45 | 7333 | 28.4% | 25.4% | 6793 | 27.3% | 24.5% |
| 46-59 | 8400 | 32.6% | 24.0% | 8153 | 32.8% | 24.0% |
| 60+ | 8235 | 31.9% | 32.7% | 8217 | 33.1% | 33.8% |
| Ethnicity, BAME | 1176 | 4.6% | 13.1% | 1104 | 4.4% | 13.2% |
| Education |  |  |  |  |  |  |
| GCSE or below | 3414 | 13.2% | 33.5% | 3202 | 12.9% | 33.0% |
| A-levels or equivalent | 4400 | 17.1% | 32.7% | 4185 | 16.8% | 32.6% |
| Degree or above | 17984 | 69.7% | 33.8% | 17451 | 70.3% | 34.4% |
| Household income <30k | 9670 | 37.5% | 47.3% | 9292 | 37.4% | 47.4% |
| Living status |  |  |  |  |  |  |
| Alone | 5182 | 20.1% | 19.3% | 5080 | 20.5% | 19.3% |
| With others (not children) | 13915 | 53.9% | 56.8% | 13476 | 54.3% | 57.7% |
| With others (including children) | 6701 | 26.0% | 23.9% | 6282 | 25.3% | 23.0% |
| Diagnosed mental illness | 4560 | 17.7% | 18.7% | 4412 | 17.8% | 19.2% |

|  | Week 7 | | | Week 8 | | |
| --- | --- | --- | --- | --- | --- | --- |
|  | Raw | | Weighted | Raw | | Weighted |
|  | N | % | % | N | % | % |
| Gender, women | 18654 | 75.6% | 50.2% | 19155 | 75.6% | 50.2% |
| Age |  |  |  |  |  |  |
| 18-29 | 1576 | 6.4% | 17.2% | 1601 | 6.3% | 16.3% |
| 30-45 | 6559 | 26.6% | 23.7% | 6564 | 25.9% | 23.3% |
| 46-59 | 8187 | 33.2% | 24.6% | 8419 | 33.2% | 25.0% |
| 60+ | 8367 | 33.9% | 34.5% | 8737 | 34.5% | 35.4% |
| Ethnicity, BAME | 1114 | 4.5% | 13.0% | 1172 | 4.6% | 13.2% |
| Education |  |  |  |  |  |  |
| GCSE or below | 3159 | 12.8% | 33.0% | 3224 | 12.7% | 32.8% |
| A-levels or equivalent | 4125 | 16.7% | 32.3% | 4196 | 16.6% | 32.1% |
| Degree or above | 17405 | 70.5% | 34.8% | 17901 | 70.7% | 35.1% |
| Household income <30k | 9196 | 37.2% | 46.7% | 9429 | 37.2% | 46.6% |
| Living status |  |  |  |  |  |  |
| Alone | 5186 | 21.0% | 19.9% | 5383 | 21.3% | 20.0% |
| With others (not children) | 13386 | 54.2% | 57.3% | 13786 | 54.4% | 57.5% |
| With others (including children) | 6117 | 24.8% | 22.8% | 6152 | 24.3% | 22.5% |
| Diagnosed mental illness | 4246 | 17.2% | 18.7% | 4348 | 17.2% | 18.7% |

|  | Week 9 | | | Week 10 | | |
| --- | --- | --- | --- | --- | --- | --- |
|  | Raw | | Weighted | Raw | | Weighted |
|  | N | % | % | N | % | % |
| Gender, women | 18548 | 75.5% | 49.9% | 17992 | 75.9% | 50.5% |
| Age |  |  |  |  |  |  |
| 18-29 | 1521 | 6.2% | 16.3% | 1411 | 5.9% | 15.5% |
| 30-45 | 6248 | 25.4% | 22.8% | 5860 | 24.7% | 22.3% |
| 46-59 | 8192 | 33.4% | 25.0% | 7985 | 33.7% | 25.4% |
| 60+ | 8592 | 35.0% | 35.9% | 8462 | 35.7% | 36.9% |
| Ethnicity, BAME | 1092 | 4.4% | 13.1% | 1050 | 4.4% | 12.9% |
| Education |  |  |  |  |  |  |
| GCSE or below | 3114 | 12.7% | 32.7% | 3017 | 12.7% | 32.8% |
| A-levels or equivalent | 4082 | 16.6% | 32.5% | 3933 | 16.6% | 32.2% |
| Degree or above | 17357 | 70.7% | 34.8% | 16768 | 70.7% | 35.0% |
| Household income <30k | 9239 | 37.6% | 46.8% | 8937 | 37.7% | 46.9% |
| Living status |  |  |  |  |  |  |
| Alone | 5238 | 21.3% | 20.2% | 5109 | 21.5% | 20.4% |
| With others (not children) | 13437 | 54.7% | 57.6% | 13034 | 55.0% | 58.0% |
| With others (including children) | 5878 | 23.9% | 22.2% | 5575 | 23.5% | 21.7% |
| Diagnosed mental illness | 4206 | 17.1% | 18.9% | 4013 | 16.9% | 18.4% |

|  | Week 11 | | | Week 12 | | |
| --- | --- | --- | --- | --- | --- | --- |
|  | Raw | | Weighted | Raw | | Weighted |
|  | N | % | % | N | % | % |
| Gender, women | 17465 | 75.9% | 50.5% | 16725 | 75.8% | 50.5% |
| Age |  |  |  |  |  |  |
| 18-29 | 1308 | 5.7% | 15.3% | 1214 | 5.5% | 14.5% |
| 30-45 | 5594 | 24.3% | 21.9% | 5219 | 23.7% | 21.3% |
| 46-59 | 7724 | 33.6% | 25.2% | 7369 | 33.4% | 25.3% |
| 60+ | 8394 | 36.5% | 37.6% | 8261 | 37.4% | 38.8% |
| Ethnicity, BAME | 984 | 4.3% | 12.4% | 907 | 4.1% | 11.9% |
| Education |  |  |  |  |  |  |
| GCSE or below | 2922 | 12.7% | 32.9% | 2852 | 12.9% | 34.0% |
| A-levels or equivalent | 3801 | 16.5% | 32.3% | 3635 | 16.5% | 31.5% |
| Degree or above | 16297 | 70.8% | 34.8% | 15576 | 70.6% | 34.5% |
| Household income <30k | 8766 | 38.1% | 47.8% | 8506 | 38.6% | 47.9% |
| Living status |  |  |  |  |  |  |
| Alone | 5042 | 21.9% | 20.5% | 4857 | 22.0% | 20.8% |
| With others (not children) | 12665 | 55.0% | 58.3% | 12211 | 55.3% | 58.6% |
| With others (including children) | 5313 | 23.1% | 21.2% | 4995 | 22.6% | 20.7% |
| Diagnosed mental illness | 3866 | 16.8% | 18.3% | 3657 | 16.6% | 18.1% |

|  | Week 13 | | | Week 14 | | |
| --- | --- | --- | --- | --- | --- | --- |
|  | Raw | | Weighted | Raw | | Weighted |
|  | N | % | % | N | % | % |
| Gender, women | 16233 | 75.7% | 49.8% | 15818 | 75.8% | 50.0% |
| Age |  |  |  |  |  |  |
| 18-29 | 1154 | 5.4% | 14.5% | 1074 | 5.1% | 14.1% |
| 30-45 | 5008 | 23.4% | 21.2% | 4806 | 23.0% | 20.7% |
| 46-59 | 7133 | 33.3% | 24.9% | 6940 | 33.2% | 25.1% |
| 60+ | 8139 | 38.0% | 39.4% | 8057 | 38.6% | 40.1% |
| Ethnicity, BAME | 892 | 4.2% | 12.2% | 857 | 4.1% | 11.9% |
| Education |  |  |  |  |  |  |
| GCSE or below | 2750 | 12.8% | 33.6% | 2733 | 13.1% | 34.2% |
| A-levels or equivalent | 3519 | 16.4% | 31.8% | 3431 | 16.4% | 31.5% |
| Degree or above | 15165 | 70.8% | 34.6% | 14713 | 70.5% | 34.3% |
| Household income <30k | 8284 | 38.6% | 48.0% | 8134 | 39.0% | 48.2% |
| Living status |  |  |  |  |  |  |
| Alone | 4758 | 22.2% | 20.9% | 4659 | 22.3% | 21.0% |
| With others (not children) | 11897 | 55.5% | 58.9% | 11612 | 55.6% | 59.1% |
| With others (including children) | 4779 | 22.3% | 20.2% | 4606 | 22.1% | 19.9% |
| Diagnosed mental illness | 3536 | 16.5% | 17.6% | 3404 | 16.3% | 17.6% |

|  | Week 15 | | | Week 16 | | |
| --- | --- | --- | --- | --- | --- | --- |
|  | Raw | | Weighted | Raw | | Weighted |
|  | N | % | % | N | % | % |
| Gender, women | 15211 | 75.5% | 49.7% | 14644 | 75.7% | 49.8% |
| Age |  |  |  |  |  |  |
| 18-29 | 968 | 4.8% | 12.9% | 922 | 4.8% | 12.9% |
| 30-45 | 4503 | 22.3% | 20.2% | 4279 | 22.1% | 19.9% |
| 46-59 | 6736 | 33.4% | 25.5% | 6411 | 33.1% | 25.0% |
| 60+ | 7942 | 39.4% | 41.4% | 7745 | 40.0% | 42.2% |
| Ethnicity, BAME | 789 | 3.9% | 11.3% | 771 | 4.0% | 11.4% |
| Education |  |  |  |  |  |  |
| GCSE or below | 2651 | 13.2% | 35.1% | 2582 | 13.3% | 35.1% |
| A-levels or equivalent | 3342 | 16.6% | 30.9% | 3193 | 16.5% | 31.2% |
| Degree or above | 14156 | 70.3% | 34.0% | 13582 | 70.2% | 33.7% |
| Household income <30k | 7912 | 39.3% | 48.4% | 7665 | 39.6% | 49.3% |
| Living status |  |  |  |  |  |  |
| Alone | 4561 | 22.6% | 21.2% | 4424 | 22.9% | 21.4% |
| With others (not children) | 11253 | 55.8% | 59.2% | 10821 | 55.9% | 59.4% |
| With others (including children) | 4335 | 21.5% | 19.6% | 4112 | 21.2% | 19.2% |
| Diagnosed mental illness | 3238 | 16.1% | 17.1% | 3154 | 16.3% | 17.3% |

|  | Week 17 | | | Week 18 | | |
| --- | --- | --- | --- | --- | --- | --- |
|  | Raw | | Weighted | Raw | | Weighted |
|  | N | % | % | N | % | % |
| Gender, women | 14272 | 75.5% | 49.4% | 13875 | 75.5% | 49.3% |
| Age |  |  |  |  |  |  |
| 18-29 | 859 | 4.5% | 12.4% | 829 | 4.5% | 12.2% |
| 30-45 | 4104 | 21.7% | 20.0% | 3925 | 21.4% | 19.7% |
| 46-59 | 6238 | 33.0% | 25.1% | 6055 | 33.0% | 25.3% |
| 60+ | 7707 | 40.8% | 42.5% | 7560 | 41.2% | 42.9% |
| Ethnicity, BAME | 764 | 4.0% | 11.5% | 735 | 4.0% | 11.2% |
| Education |  |  |  |  |  |  |
| GCSE or below | 2535 | 13.4% | 35.3% | 2496 | 13.6% | 35.6% |
| A-levels or equivalent | 3148 | 16.6% | 31.4% | 3061 | 16.7% | 31.1% |
| Degree or above | 13225 | 69.9% | 33.4% | 12812 | 69.7% | 33.3% |
| Household income <30k | 7525 | 39.8% | 49.5% | 7381 | 40.2% | 49.5% |
| Living status |  |  |  |  |  |  |
| Alone | 4307 | 22.8% | 21.6% | 4227 | 23.0% | 21.9% |
| With others (not children) | 10628 | 56.2% | 59.5% | 10335 | 56.3% | 59.5% |
| With others (including children) | 3973 | 21.0% | 18.9% | 3807 | 20.7% | 18.5% |
| Diagnosed mental illness | 3021 | 16.0% | 17.0% | 2933 | 16.0% | 16.9% |

|  | Week 19 | | | Week 20 | | |
| --- | --- | --- | --- | --- | --- | --- |
|  | Raw | | Weighted | Raw | | Weighted |
|  | N | % | % | N | % | % |
| Gender, women | 13341 | 75.5% | 49.2% | 12948 | 75.5% | 49.6% |
| Age |  |  |  |  |  |  |
| 18-29 | 790 | 4.5% | 12.5% | 739 | 4.3% | 11.7% |
| 30-45 | 3741 | 21.2% | 19.6% | 3525 | 20.6% | 19.1% |
| 46-59 | 5791 | 32.8% | 24.9% | 5656 | 33.0% | 25.4% |
| 60+ | 7351 | 41.6% | 43.0% | 7219 | 42.1% | 43.9% |
| Ethnicity, BAME | 696 | 3.9% | 11.6% | 665 | 3.9% | 10.8% |
| Education |  |  |  |  |  |  |
| GCSE or below | 2372 | 13.4% | 35.2% | 2347 | 13.7% | 35.5% |
| A-levels or equivalent | 2956 | 16.7% | 31.6% | 2866 | 16.7% | 31.5% |
| Degree or above | 12345 | 69.9% | 33.2% | 11926 | 69.6% | 33.0% |
| Household income <30k | 7108 | 40.2% | 49.4% | 6920 | 40.4% | 49.8% |
| Living status |  |  |  |  |  |  |
| Alone | 4114 | 23.3% | 21.8% | 3969 | 23.2% | 22.2% |
| With others (not children) | 10006 | 56.6% | 59.9% | 9750 | 56.9% | 59.6% |
| With others (including children) | 3553 | 20.1% | 18.3% | 3420 | 20.0% | 18.2% |
| Diagnosed mental illness | 2819 | 16.0% | 16.9% | 2710 | 15.8% | 16.9% |

Table S5 Categorisation of the severity of anxiety and depressive symptoms week by week

|  | GAD | | | | | | | | PHQ | | | | | | | | | |
| --- | --- | --- | --- | --- | --- | --- | --- | --- | --- | --- | --- | --- | --- | --- | --- | --- | --- | --- |
|  | 0-4 | | 5-9 | | 10-14 | | 15+ | | 0-4 | | 5-9 | | 10-14 | | 15-19 | | 20+ | |
| Week | % | N | % | N | % | N | % | N | % | N | % | N | % | N | % | N | % | N |
| 1 | 53.4% | 9123 | 24.1% | 4111 | 12.2% | 2092 | 10.3% | 1764 | 48.1% | 8228 | 26.8% | 4578 | 13.0% | 2218 | 7.5% | 1290 | 4.5% | 776 |
| 2 | 55.0% | 10681 | 24.0% | 4657 | 12.4% | 2405 | 8.7% | 1681 | 44.9% | 8719 | 28.3% | 5502 | 15.1% | 2933 | 7.6% | 1468 | 4.1% | 803 |
| 3 | 57.7% | 15295 | 22.9% | 6085 | 11.3% | 2996 | 8.1% | 2142 | 45.9% | 12175 | 27.5% | 7292 | 14.8% | 3928 | 7.3% | 1942 | 4.5% | 1180 |
| 4 | 58.7% | 15580 | 23.1% | 6122 | 10.6% | 2807 | 7.6% | 2028 | 46.9% | 12458 | 27.6% | 7316 | 14.4% | 3821 | 6.8% | 1810 | 4.3% | 1132 |
| 5 | 59.8% | 15428 | 23.0% | 5947 | 9.9% | 2550 | 7.3% | 1879 | 48.9% | 12612 | 27.3% | 7049 | 12.9% | 3337 | 6.8% | 1758 | 4.1% | 1047 |
| 6 | 60.0% | 14461 | 22.1% | 5330 | 10.0% | 2417 | 7.8% | 1876 | 46.0% | 11069 | 26.8% | 6458 | 14.9% | 3594 | 7.1% | 1707 | 5.2% | 1256 |
| 7 | 60.9% | 14358 | 21.6% | 5088 | 10.7% | 2516 | 6.8% | 1596 | 48.0% | 11305 | 25.6% | 6030 | 14.1% | 3314 | 7.6% | 1783 | 4.8% | 1125 |
| 8 | 62.3% | 14914 | 21.0% | 5025 | 9.7% | 2312 | 7.0% | 1681 | 49.1% | 11756 | 25.9% | 6196 | 13.1% | 3140 | 7.3% | 1739 | 4.6% | 1102 |
| 9 | 63.2% | 14632 | 20.0% | 4622 | 10.0% | 2317 | 6.8% | 1579 | 50.4% | 11674 | 24.8% | 5737 | 13.1% | 3039 | 6.8% | 1579 | 4.8% | 1123 |
| 10 | 65.1% | 14299 | 19.7% | 4337 | 8.5% | 1873 | 6.7% | 1462 | 53.4% | 11726 | 24.5% | 5372 | 12.0% | 2633 | 6.0% | 1316 | 4.2% | 922 |
| 11 | 65.7% | 13874 | 19.2% | 4061 | 8.4% | 1773 | 6.6% | 1395 | 54.2% | 11430 | 23.9% | 5037 | 12.0% | 2523 | 5.5% | 1165 | 4.5% | 947 |
| 12 | 65.1% | 13129 | 19.7% | 3979 | 9.0% | 1805 | 6.2% | 1240 | 52.6% | 10610 | 24.5% | 4942 | 12.1% | 2443 | 6.0% | 1211 | 4.7% | 946 |
| 13 | 67.1% | 13147 | 18.4% | 3603 | 8.5% | 1662 | 6.0% | 1169 | 54.4% | 10655 | 24.6% | 4814 | 11.0% | 2164 | 5.6% | 1094 | 4.4% | 855 |
| 14 | 68.3% | 12982 | 18.4% | 3488 | 8.1% | 1543 | 5.2% | 986 | 57.3% | 10879 | 23.5% | 4459 | 10.3% | 1957 | 5.3% | 1016 | 3.6% | 686 |
| 15 | 69.1% | 12565 | 17.5% | 3188 | 8.2% | 1496 | 5.1% | 936 | 57.1% | 10391 | 23.8% | 4322 | 10.2% | 1862 | 5.0% | 913 | 3.8% | 697 |
| 16 | 69.3% | 12159 | 18.3% | 3212 | 7.1% | 1241 | 5.3% | 934 | 58.3% | 10222 | 23.4% | 4103 | 9.8% | 1727 | 4.9% | 851 | 3.7% | 642 |
| 17 | 69.9% | 11987 | 17.9% | 3062 | 7.1% | 1218 | 5.1% | 880 | 60.9% | 10436 | 21.7% | 3725 | 8.9% | 1524 | 5.0% | 855 | 3.5% | 607 |
| 18 | 71.7% | 11946 | 16.3% | 2720 | 6.8% | 1130 | 5.2% | 858 | 62.0% | 10325 | 21.0% | 3503 | 8.9% | 1487 | 4.6% | 760 | 3.5% | 579 |
| 19 | 71.6% | 11475 | 16.9% | 2715 | 6.6% | 1058 | 4.9% | 778 | 62.4% | 9993 | 21.3% | 3416 | 8.6% | 1384 | 4.5% | 726 | 3.2% | 507 |
| 20 | 71.7% | 11068 | 16.8% | 2586 | 6.5% | 996 | 5.0% | 779 | 63.3% | 9769 | 20.3% | 3134 | 8.4% | 1296 | 4.5% | 699 | 3.4% | 529 |

Table S6 Results from the conditional multi-process LGM model

|  | **Anxiety** | | | **Depression** | | |
| --- | --- | --- | --- | --- | --- | --- |
|  | b | se | p | b | se | p |
| **Main Structure** |  |  |  |  |  |  |
| Intercept (I) | 5.35 | 0.23 | <0.0001 | 7.49 | 0.25 | <0.0001 |
| Slope (S) | -1.93 | 0.26 | <0.0001 | -2.52 | 0.28 | <0.0001 |
| **Slope Structure** |  |  |  |  |  |  |
| Week 1 (Y_0_) | 0.00 | -- | -- | 0.00 | -- | -- |
| Week 2 (Y_1_) | 0.04 | 0.02 | 0.020 | -0.03 | 0.02 | 0.171 |
| Week 3 (Y_1_) | 0.17 | 0.02 | <0.0001 | 0.07 | 0.02 | 0.004 |
| Week 4 (Y_1_) | 0.28 | 0.02 | <0.0001 | 0.19 | 0.02 | <0.0001 |
| Week 5 (Y_1_) | 0.36 | 0.02 | <0.0001 | 0.29 | 0.02 | <0.0001 |
| Week 6 (Y_1_) | 0.39 | 0.02 | <0.0001 | 0.26 | 0.02 | <0.0001 |
| Week 7 (Y_1_) | 0.46 | 0.02 | <0.0001 | 0.33 | 0.02 | <0.0001 |
| Week 8 (Y_1_) | 0.53 | 0.02 | <0.0001 | 0.41 | 0.02 | <0.0001 |
| Week 9 (Y_1_) | 0.60 | 0.02 | <0.0001 | 0.49 | 0.02 | <0.0001 |
| Week 10 (Y_1_) | 0.68 | 0.02 | <0.0001 | 0.60 | 0.02 | <0.0001 |
| Week 11 (Y_1_) | 0.72 | 0.02 | <0.0001 | 0.64 | 0.03 | <0.0001 |
| Week 12 (Y_1_) | 0.75 | 0.02 | <0.0001 | 0.68 | 0.03 | <0.0001 |
| Week 13 (Y_1_) | 0.82 | 0.02 | <0.0001 | 0.76 | 0.02 | <0.0001 |
| Week 14 (Y_1_) | 0.89 | 0.02 | <0.0001 | 0.85 | 0.02 | <0.0001 |
| Week 15 (Y_1_) | 0.93 | 0.02 | <0.0001 | 0.89 | 0.02 | <0.0001 |
| Week 16 (Y_1_) | 0.96 | 0.02 | <0.0001 | 0.94 | 0.02 | <0.0001 |
| Week 17 (Y_1_) | 0.98 | 0.02 | <0.0001 | 1.00 | 0.02 | <0.0001 |
| Week 18 (Y_1_) | 1.01 | 0.02 | <0.0001 | 0.99 | 0.02 | <0.0001 |
| Week 19 (Y_1_) | 1.00 | 0.02 | <0.0001 | 1.00 | 0.02 | <0.0001 |
| Week 20 (Y_1_) | 1.00 | -- | -- | 1.00 | -- | -- |
| **Covariance Structure** |  | |  | |  | |
| I (anxiety) with S (anxiety) | -9.40 | | 0.71 | | <0.0001 | |
| I (depression) with S (depression) | -6.99 | | 0.73 | | <0.0001 | |
| I (anxiety) with I (depression) | 20.46 | | 0.56 | | <0.0001 | |
| S (anxiety) with S (depression) | 18.64 | | 1.23 | | <0.0001 | |
| I (anxiety) with S (depression) | -8.53 | | 0.73 | | <0.0001 | |
| I (depression) with S (anxiety) | -7.07 | | 0.68 | | <0.0001 | |

Notes: Model controlled for covariates, including gender, age, ethnicity, education, income, living status and pre-existing mental health conditions
